# Half-Dose Ticagrelor Monotherapy Versus Standard Dual Antiplatelet Therapy in Chronic Coronary Syndrome After Percutaneous Coronary Intervention: A Randomized Pilot Trial With PRU-Guided Pharmacodynamic Assessment

**DOI:** 10.64898/2026.06.29.26356433

**Authors:** Feng-Yu Kuo, Mei-Chi Wang, Cheng-Hung Chiang, En-Shao Liu, Tse-Hsuan Yang, Haw-Ting Tai, Cai-Sin Yao, Ren-In Chang, Guan Yan Mar

## Abstract

**Background:** Aspirin-free P2Y_12_-inhibitor monotherapy after percutaneous coronary intervention (PCI) is an alternative to dual antiplatelet therapy (DAPT), but the evidence rests largely on full-dose ticagrelor in acute coronary syndrome and on designs retaining a DAPT run-in; East-Asian patients may not require the same antithrombotic intensity. We compared standard DAPT, DAPT with half-dose ticagrelor, and aspirin-free half-dose ticagrelor monotherapy initiated on the day of PCI in chronic coronary syndrome (CCS).

**Methods:** Sixty-one East-Asian patients with CCS scheduled for elective PCI were randomized 1:1:1 to Control (aspirin plus clopidogrel), Experimental A (aspirin plus ticagrelor 45 mg twice daily), or Experimental B (ticagrelor 45 mg monotherapy, aspirin discontinued at day 2). DAPT arms continued for six months; Experimental B continued indefinitely. P2Y_12_ reaction units (PRU) were measured at baseline and at a median of 17 days.

**Results:** PRU reduction was three-fold greater in both ticagrelor arms than in Control (ΔPRU −188 and −181 versus −60.5; *P*<0.001), with no difference between ticagrelor arms (*P*=0.772). At 12 months, major adverse cardiovascular events (MACE) and clinically relevant bleeding each occurred in 1 of 17 Experimental B patients (5.9%) and in neither other arm. One Experimental A patient crossed over for ticagrelor-induced dyspnea; no stent thrombosis or cardiac death occurred.

**Conclusions:** In East-Asian patients with CCS, half-dose ticagrelor produced markedly greater platelet inhibition than standard DAPT, with an identical effect whether given with or without aspirin. It merits evaluation in an adequately powered randomized trial.

**Clinical Trial Registration:** URL: https://www.clinicaltrials.gov; Unique Identifier: NCT07622056.

**Clinical Perspective:**

- What Is New? This three-arm randomized pilot trial is the first to demonstrate that aspirin-free half-dose ticagrelor monotherapy (45 mg twice daily) initiated on the day of percutaneous coronary intervention — without any dual antiplatelet run-in period — produces platelet inhibition approximately three-fold greater than standard aspirin-plus-clopidogrel therapy in East-Asian patients with chronic coronary syndrome, with an identical pharmacodynamic effect whether aspirin is retained or omitted.
- What Are the Clinical Implications? For East-Asian patients with chronic coronary syndrome undergoing elective percutaneous coronary intervention, a simplified aspirin-free half-dose ticagrelor monotherapy strategy initiated at the index procedure achieves superior platelet inhibition with a favorable 12-month safety profile — no stent thrombosis, no cardiac death, and only one non-major gastrointestinal bleed — supporting the feasibility of a definitive adequately powered randomized trial to validate this approach and potentially reduce bleeding risk without compromising ischemic protection in this East-Asian population.

## Introduction

Six months of aspirin plus clopidogrel remains the guideline-recommended antiplatelet regimen after PCI for CCS,^1^ with the more potent P2Y_12_ inhibitors reserved for the acute coronary syndromes (ACS) that generated their pivotal evidence.^2,3^ Yet the contemporary landscape of post-PCI antithrombotic therapy has shifted considerably since PLATO and TRITON-TIMI 38, and the conventional algorithm sits increasingly at odds with two well-established realities.

The first is the consistent demonstration that aspirin contributes more to bleeding than to ischemic protection in patients adequately treated with a P2Y_12_ inhibitor. TWILIGHT,^4^ TICO,^5^ T-PASS,^6^ ULTIMATE-DAPT,^7^ and GLOBAL LEADERS^8^ have, in sequence, established that withdrawing aspirin after a defined period of P2Y_12_-based DAPT reduces clinically relevant bleeding without an excess of ischemic events. Each of these trials, however, retained ticagrelor at the standard 90 mg twice-daily dose, required a run-in period of conventional DAPT before aspirin was stopped, and enrolled populations dominated by ACS.

The second is the persistent and mechanistically grounded observation that East-Asian patients tolerate less antithrombotic intensity than the Caucasian populations on whom dose recommendations were calibrated.^9–11^ Ticagrelor exposure is higher in East-Asian patients, baseline platelet reactivity is lower, and the bleeding-to-ischemia ratio is shifted relative to PLATO-era expectations. Pharmacokinetic and pharmacodynamic studies in Chinese and Japanese cohorts have shown that ticagrelor 45 mg twice daily — half the standard maintenance dose — yields platelet inhibition statistically indistinguishable from the 90 mg regimen.^12–14^

No randomized trial has yet tested an aspirin-free, half-dose ticagrelor monotherapy strategy initiated at the index PCI in CCS patients. The closest registered protocol of which we are aware (NCT07080684) uses ticagrelor 60 mg twice daily following a one-month DAPT lead-in, with a two-arm design. We therefore conducted a three-arm randomized pilot trial in East-Asian CCS patients to compare standard DAPT, DAPT with half-dose ticagrelor, and aspirin-free half-dose ticagrelor monotherapy from the day of PCI, using the change in PRU as the primary pharmacodynamic readout and 12-month clinical events as exploratory endpoints.

## Methods

### Study design and ethics

This was a prospective, single-center, randomized, open-label, parallel-group pilot trial conducted at Kaohsiung Veterans General Hospital from 1 January 2024 to 31 December 2024. The protocol was approved by the institutional review board (KSVGH 23-CT12-07), and the trial was conducted in accordance with the Declaration of Helsinki. Written informed consent was obtained from every participant before randomization. The trial was registered at ClinicalTrials.gov (NCT07622056); registration was completed retrospectively.

### Patients

Eligible patients were adults with CCS, defined per the 2019 ESC criteria,¹ scheduled for elective PCI on the day after admission. Exclusion criteria were: ACS at presentation; prior intolerance to any study drug; active bleeding or BARC ≥ 3 bleeding within three months;^15^ NYHA class IV heart failure; platelet count below 100 × 10^9^/L; eGFR below 30 mL/min/1.73 m^2^; severe hepatic dysfunction; planned non-cardiac surgery within 12 months; or inability to provide informed consent. Patients in whom PCI was not performed at the index procedure were withdrawn from the per-protocol analysis.

### Randomization and interventions

Patients were randomized 1:1:1 by computer-generated sequence. Treatment allocation was open-label; PRU assays were performed by laboratory personnel blinded to group assignment. Loading doses were administered after randomization and before PCI on the following day.

- Control (aspirin plus clopidogrel): aspirin 100 mg plus a 300 mg clopidogrel loading dose; then aspirin 100 mg plus clopidogrel 75 mg once daily for six months, followed by aspirin 100 mg monotherapy.
- Experimental A (aspirin plus half-dose ticagrelor): aspirin 100 mg plus a 180 mg ticagrelor loading dose; then aspirin 100 mg plus ticagrelor 45 mg twice daily for six months, followed by aspirin 100 mg monotherapy.
- Experimental B (half-dose ticagrelor monotherapy): aspirin 100 mg plus a 180 mg ticagrelor loading dose on the day of randomization; then ticagrelor 45 mg twice daily monotherapy with aspirin discontinued at day 2 after randomization, continued indefinitely.

All procedures used contemporary second-generation drug-eluting stents and were performed by experienced operators. Procedural anticoagulation, intracoronary imaging, and physiological assessment were at operator discretion. Lesion complexity was classified according to ACC/AHA lesion morphology criteria. The number of diseased vessels, presence of chronic total occlusion, number of stents, total stent length, and use of intravascular imaging (intravascular ultrasound or optical coherence tomography) were recorded for each patient. In patients with multivessel disease, the choice between surgical and percutaneous revascularization followed heart-team evaluation; those who declined coronary artery bypass grafting after detailed shared decision-making underwent complete percutaneous revascularization, which could increase stent count and total stented length. Statin therapy and other guideline-directed medical therapies were prescribed per standard care.

### Endpoints

The primary endpoint was the change in PRU from baseline to follow-up, with follow-up measured approximately two weeks after PCI. Secondary endpoints, all collected over 12 months, were: major adverse cardiovascular events (MACE — a composite of all-cause death, target-vessel or target-lesion revascularization, and recurrent myocardial infarction); any clinically relevant bleeding, classified by the Bleeding Academic Research Consortium (BARC) criteria;¹⁵ and study-drug intolerance or crossover, including ticagrelor-related dyspnea.

### Pharmacodynamic assessment

Platelet reactivity was assessed with the VerifyNow P2Y₁₂ point-of-care assay on whole-blood samples drawn at two time points: before the index loading dose, and at a median of 17 days after PCI. Results are expressed in PRU. High platelet reactivity (HPR) was pre-specified as PRU ≥ 208 and very low reactivity as PRU < 85.

### Statistical analysis

As an exploratory pilot trial, no formal sample-size calculation was performed; planned enrollment of approximately 60 patients was driven by feasibility and the explicit intent of generating effect-size estimates for a definitive trial. Continuous variables are presented as mean ± SD when normally distributed and as median (interquartile range, IQR) otherwise; categorical variables are reported as count (percentage). Between-group comparisons used one-way ANOVA or the Kruskal–Wallis test for continuous variables and Pearson χ^2^ or Fisher’s exact test for categorical variables. Within-group comparisons used the paired *t*-test or Wilcoxon signed-rank test. Post-hoc pairwise testing was Bonferroni-corrected. Time-to-event analyses used the Kaplan–Meier estimator with log-rank testing. Two-sided *P*-values below 0.05 were considered statistically significant. All analyses were performed in R version 4.3. The corresponding author had full access to all the data in the study and takes responsibility for its integrity and the data analysis.

## Results

### Patient characteristics

Sixty-one patients were enrolled and randomized: 20 to Control, 21 to Experimental A, and 20 to Experimental B. Seven patients (2, 2, and 3, respectively) did not undergo PCI at the index procedure and were withdrawn per protocol, leaving 54 in the analysis population: 18 Control, 19 Experimental A, and 17 Experimental B (Figure 1). The three arms were well balanced for age, sex, anthropometrics, comorbidity profile, cardiovascular history, smoking status, and background medication (Table 1). The cohort was representative of an East-Asian CCS PCI population: mean age 63.6 years, 86.8% male, with hypertension, diabetes, and dyslipidemia each present in more than half of patients. Baseline laboratory variables — including hs-CRP, lipoprotein(a), renal function, HbA1c, lipid panel, and complete blood count parameters — did not differ between arms (Table 2).

**Figure 1.**
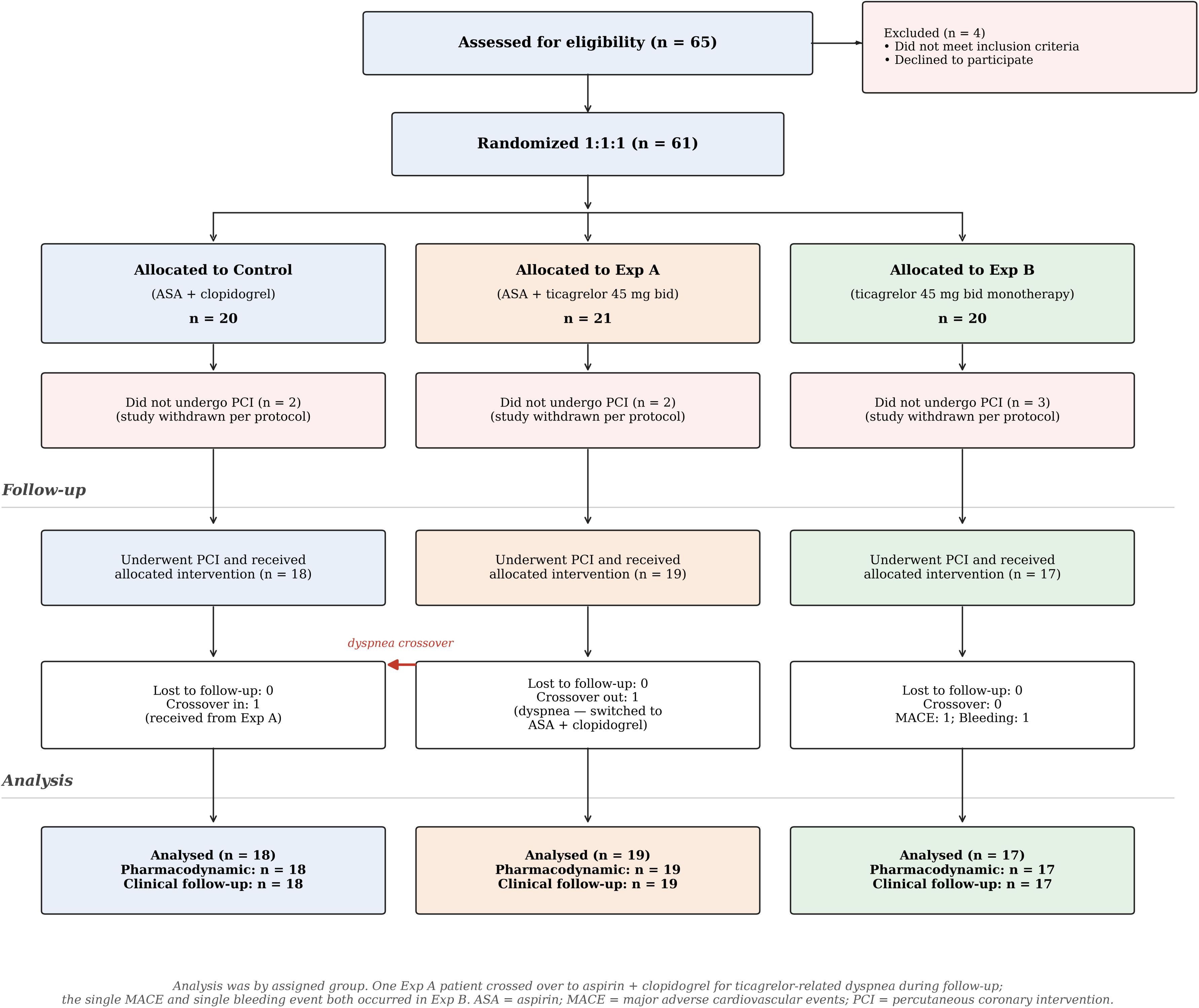
Trial flow diagram. Consort-style flow of patient screening, enrollment, randomization, and follow-up. CCS indicates chronic coronary syndrome; DAPT, dual antiplatelet therapy; PCI, percutaneous coronary intervention.

**Table 1.**
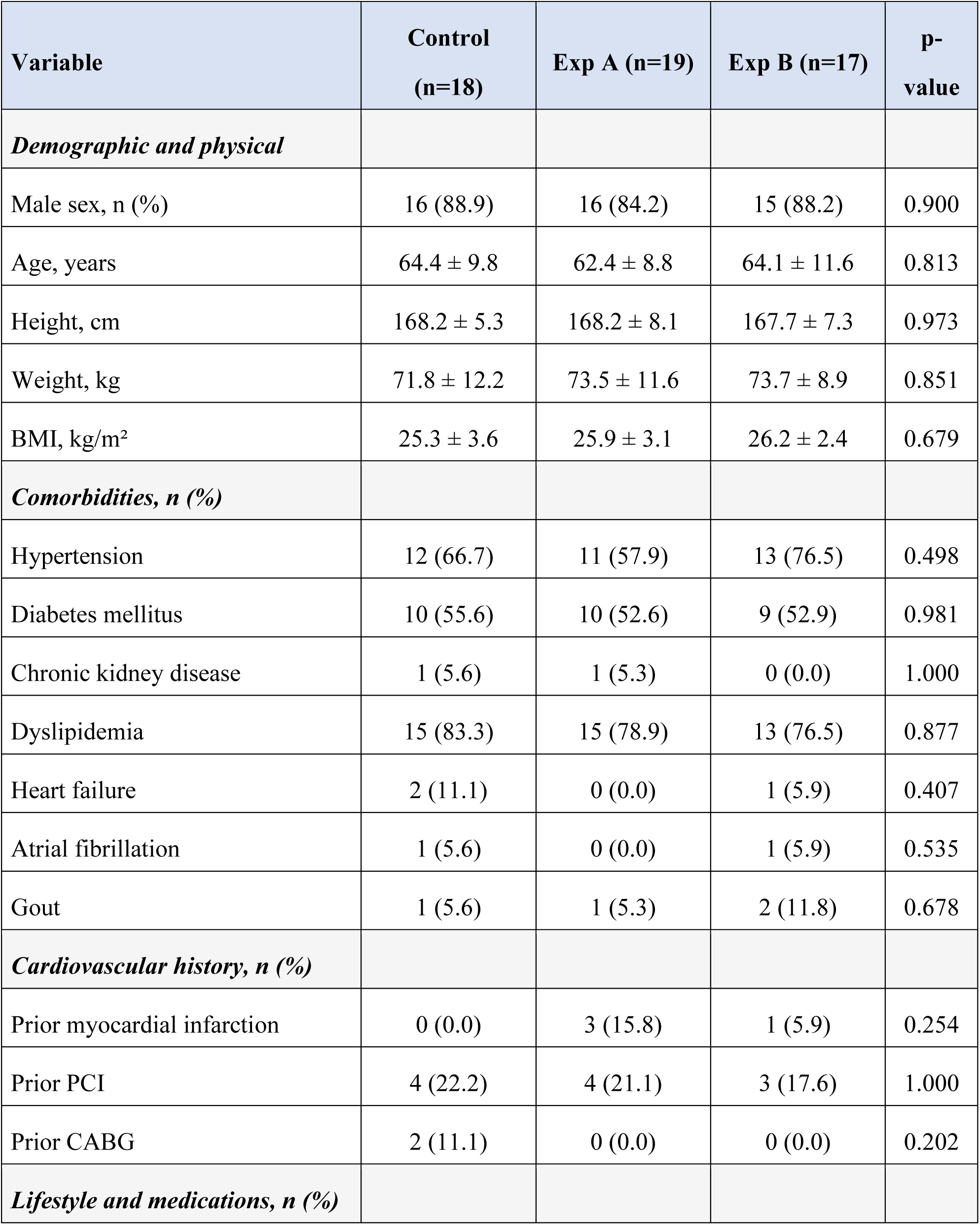
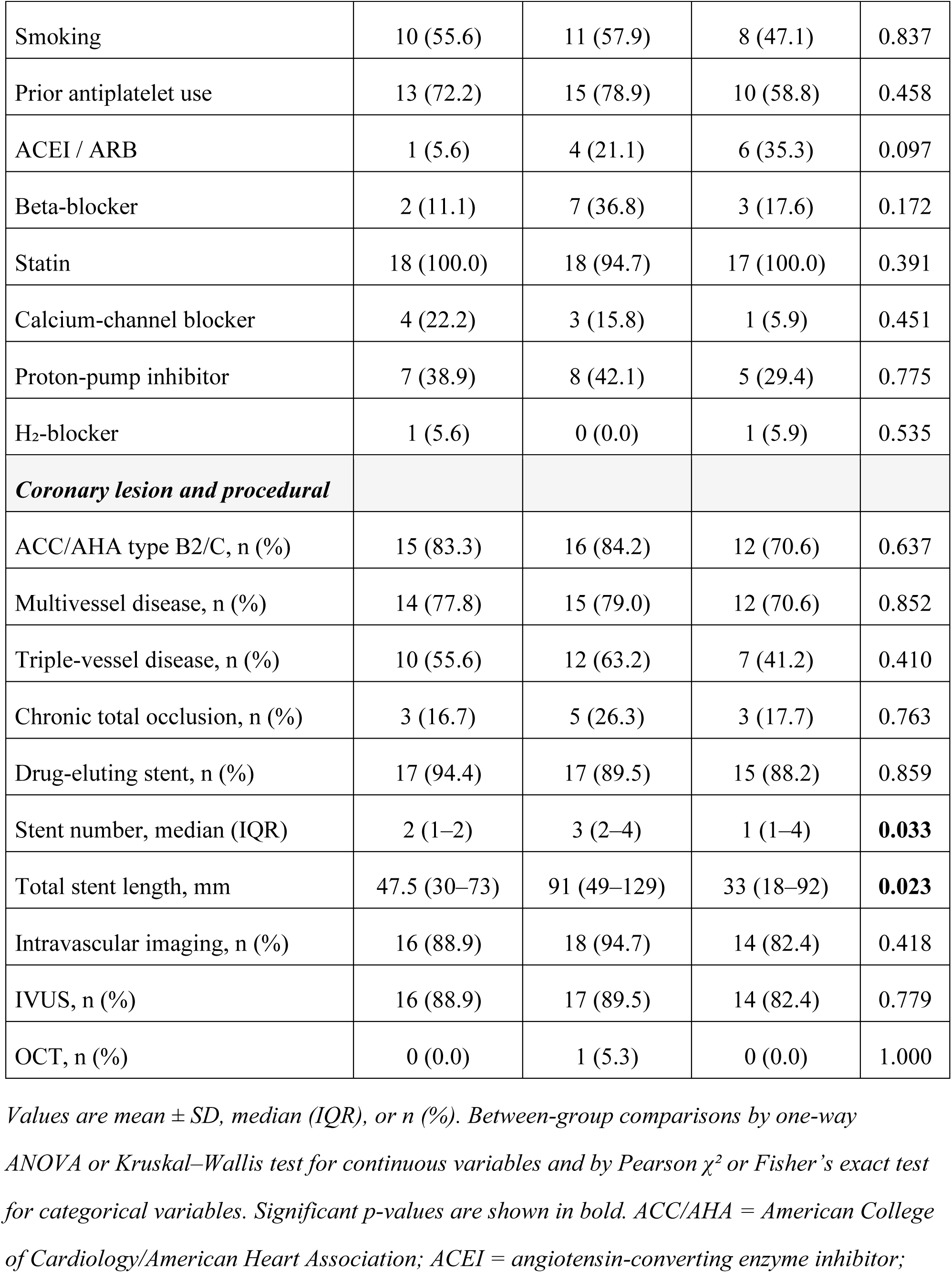
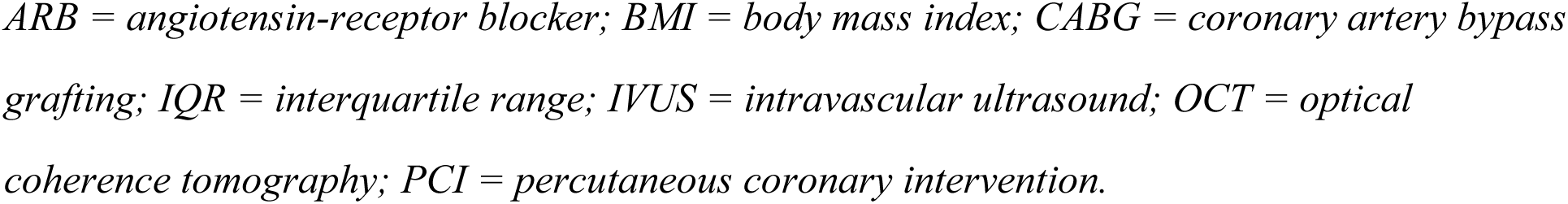
Baseline demographic, clinical, and procedural characteristics.

| **Variable** | **Control**<br>**(n=18)** | **Exp A (n=19)** | **Exp B (n=17)** | **p-**<br>**value** |
| --- | --- | --- | --- | --- |
| ***Demographic and physical*** |  |  |  |  |
| Male sex, n (%) | 16 (88.9) | 16 (84.2) | 15 (88.2) | 0.900 |
| Age, years | 64.4 ± 9.8 | 62.4 ± 8.8 | 64.1 ± 11.6 | 0.813 |
| Height, cm | 168.2 ± 5.3 | 168.2 ± 8.1 | 167.7 ± 7.3 | 0.973 |
| Weight, kg | 71.8 ± 12.2 | 73.5 ± 11.6 | 73.7 ± 8.9 | 0.851 |
| BMI, kg/m<sup>2</sup> | 25.3 ± 3.6 | 25.9 ± 3.1 | 26.2 ± 2.4 | 0.679 |
| ***Comorbidities, n (%)*** |  |  |  |  |
| Hypertension | 12 (66.7) | 11 (57.9) | 13 (76.5) | 0.498 |
| Diabetes mellitus | 10 (55.6) | 10 (52.6) | 9 (52.9) | 0.981 |
| Chronic kidney disease | 1 (5.6) | 1 (5.3) | 0 (0.0) | 1.000 |
| Dyslipidemia | 15 (83.3) | 15 (78.9) | 13 (76.5) | 0.877 |
| Heart failure | 2 (11.1) | 0 (0.0) | 1 (5.9) | 0.407 |
| Atrial fibrillation | 1 (5.6) | 0 (0.0) | 1 (5.9) | 0.535 |
| Gout | 1 (5.6) | 1 (5.3) | 2 (11.8) | 0.678 |
| ***Cardiovascular history, n (%)*** |  |  |  |  |
| Prior myocardial infarction | 0 (0.0) | 3 (15.8) | 1 (5.9) | 0.254 |
| Prior PCI | 4 (22.2) | 4 (21.1) | 3 (17.6) | 1.000 |
| Prior CABG | 2 (11.1) | 0 (0.0) | 0 (0.0) | 0.202 |
| ***Lifestyle and medications, n (%)*** |  |  |  |  |
| Smoking | 10 (55.6) | 11 (57.9) | 8 (47.1) | 0.837 |
| Prior antiplatelet use | 13 (72.2) | 15 (78.9) | 10 (58.8) | 0.458 |
| ACEI / ARB | 1 (5.6) | 4 (21.1) | 6 (35.3) | 0.097 |
| Beta-blocker | 2 (11.1) | 7 (36.8) | 3 (17.6) | 0.172 |
| Statin | 18 (100.0) | 18 (94.7) | 17 (100.0) | 0.391 |
| Calcium-channel blocker | 4 (22.2) | 3 (15.8) | 1 (5.9) | 0.451 |
| Proton-pump inhibitor | 7 (38.9) | 8 (42.1) | 5 (29.4) | 0.775 |
| H<sub>2</sub>-blocker | 1 (5.6) | 0 (0.0) | 1 (5.9) | 0.535 |
| ***Coronary lesion and procedural*** |  |  |  |  |
| ACC/AHA type B2/C, n (%) | 15 (83.3) | 16 (84.2) | 12 (70.6) | 0.637 |
| Multivessel disease, n (%) | 14 (77.8) | 15 (79.0) | 12 (70.6) | 0.852 |
| Triple-vessel disease, n (%) | 10 (55.6) | 12 (63.2) | 7 (41.2) | 0.410 |
| Chronic total occlusion, n (%) | 3 (16.7) | 5 (26.3) | 3 (17.7) | 0.763 |
| Drug-eluting stent, n (%) | 17 (94.4) | 17 (89.5) | 15 (88.2) | 0.859 |
| Stent number, median (IQR) | 2 (1–2) | 3 (2–4) | 1 (1–4) | **0.033** |
| Total stent length, mm | 47.5 (30–73) | 91 (49–129) | 33 (18–92) | **0.023** |
| Intravascular imaging, n (%) | 16 (88.9) | 18 (94.7) | 14 (82.4) | 0.418 |
| IVUS, n (%) | 16 (88.9) | 17 (89.5) | 14 (82.4) | 0.779 |
| OCT, n (%) | 0 (0.0) | 1 (5.3) | 0 (0.0) | 1.000 |

*Values are mean ± SD, median (IQR), or n (%). Between-group comparisons by one-way ANOVA or Kruskal–Wallis test for continuous variables and by Pearson $\chi^2$ or Fisher’s exact test for categorical variables. Significant p-values are shown in bold. ACC/AHA = American College of Cardiology/American Heart Association; ACEI = angiotensin-converting enzyme inhibitor;*
*ARB = angiotensin-receptor blocker; BMI = body mass index; CABG = coronary artery bypass grafting; IQR = interquartile range; IVUS = intravascular ultrasound; OCT = optical coherence tomography; PCI = percutaneous coronary intervention.*

**Table 2.**
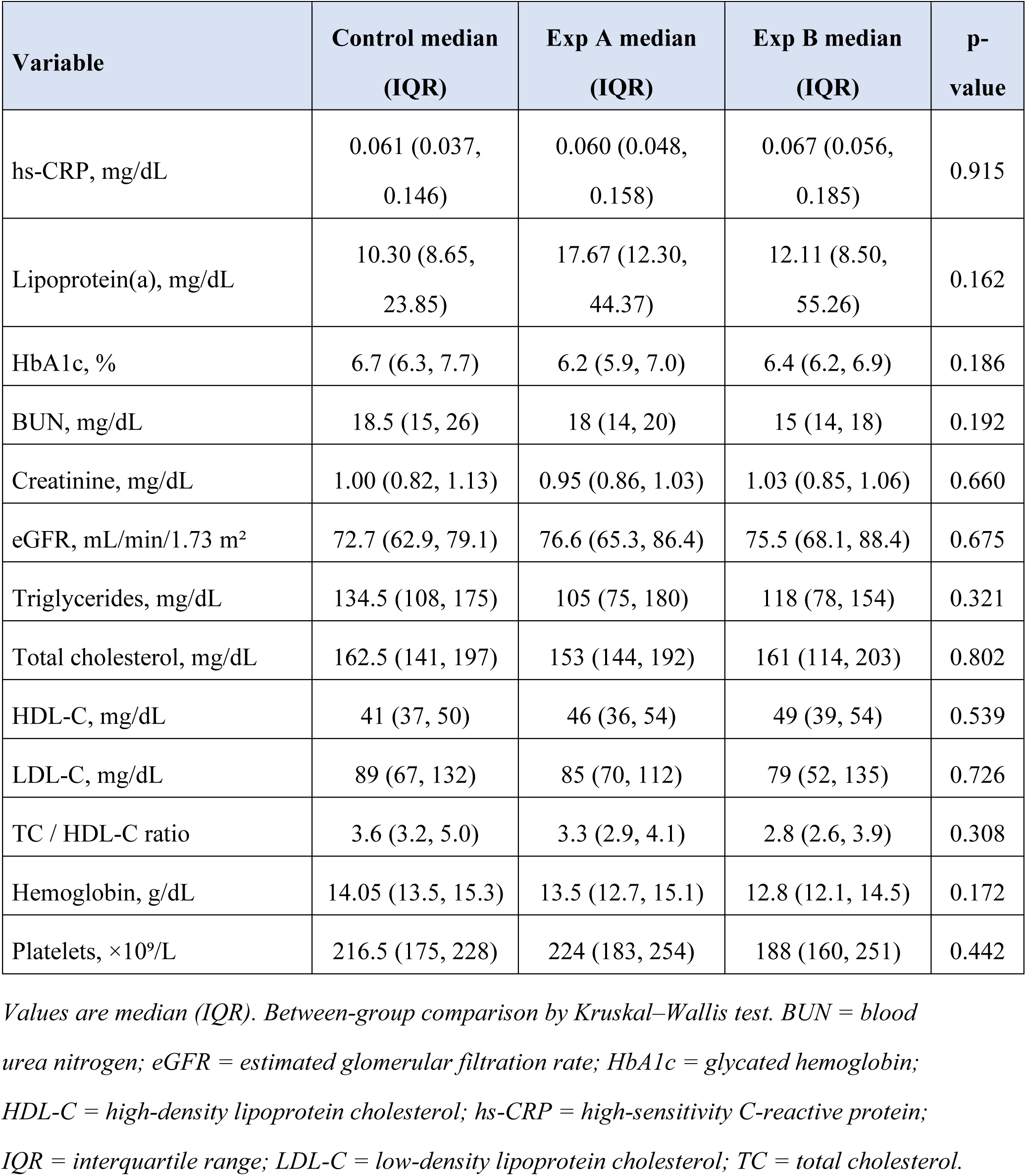
Baseline laboratory profiles.

### Coronary lesion and procedural characteristics

Lesion complexity was high and broadly comparable across arms (Table 1). ACC/AHA type B2/C lesions were present in 83.3%, 84.2%, and 70.6% of Control, Experimental A, and Experimental B patients, respectively (*P*=0.637), and multivessel disease in 77.8%, 78.9%, and 70.6% (*P*=0.852). Triple-vessel disease (55.6%, 63.2%, 41.2%; *P*=0.410) and chronic total occlusion (16.7%, 26.3%, 17.6%; *P*=0.763) did not differ significantly between groups. Drug-eluting stents were used in the great majority of patients in every arm, and intravascular imaging guided the procedure in most cases (88.9%, 94.7%, and 82.4%; *P*=0.418), predominantly by intravascular ultrasound; optical coherence tomography was used in a single Experimental A patient.

Despite pharmacodynamic equivalence between the two ticagrelor arms, they differed in procedural burden: Experimental A patients received significantly more stents (median 3 [IQR 2–4] vs. 2 [1–2] in Control and 1 [1–4] in Experimental B; *P*=0.033) and a greater total stent length (91 [49–129] mm vs. 47.5 [30–73] mm and 33 [18–92] mm; *P*=0.023). This imbalance — an expected consequence of small-sample randomization rather than selection bias — is considered further in the Discussion as a potential confounder of the clinical-event comparison.

### Pharmacodynamic outcomes

Baseline PRU was comparable across arms (medians: 211 in Control, 210 in Experimental A, 253 in Experimental B). At a median follow-up interval of 17 days (with no significant between-group difference in interval; Table 3), PRU fell significantly in every arm, but the magnitude of reduction differed markedly between strategies (Figure 2). Follow-up PRU was 167 (IQR 127–194) in Control, 38 (9–58) in Experimental A, and 53 (28–105) in Experimental B. Between-group comparison of ΔPRU (Table 3 and Figure 3) showed values of −60.5 (−76, −25) in Control, −188 (−201, −154) in Experimental A, and −181 (−227, −143) in Experimental B (overall *P*<0.001). Post-hoc pairwise testing confirmed that both ticagrelor arms produced significantly greater PRU reduction than Control (*P*<0.001 for each), whereas the two ticagrelor arms did not differ from each other (*P*=0.772). Changes in blood pressure and serum uric acid did not differ significantly between arms.

**Figure 2.**
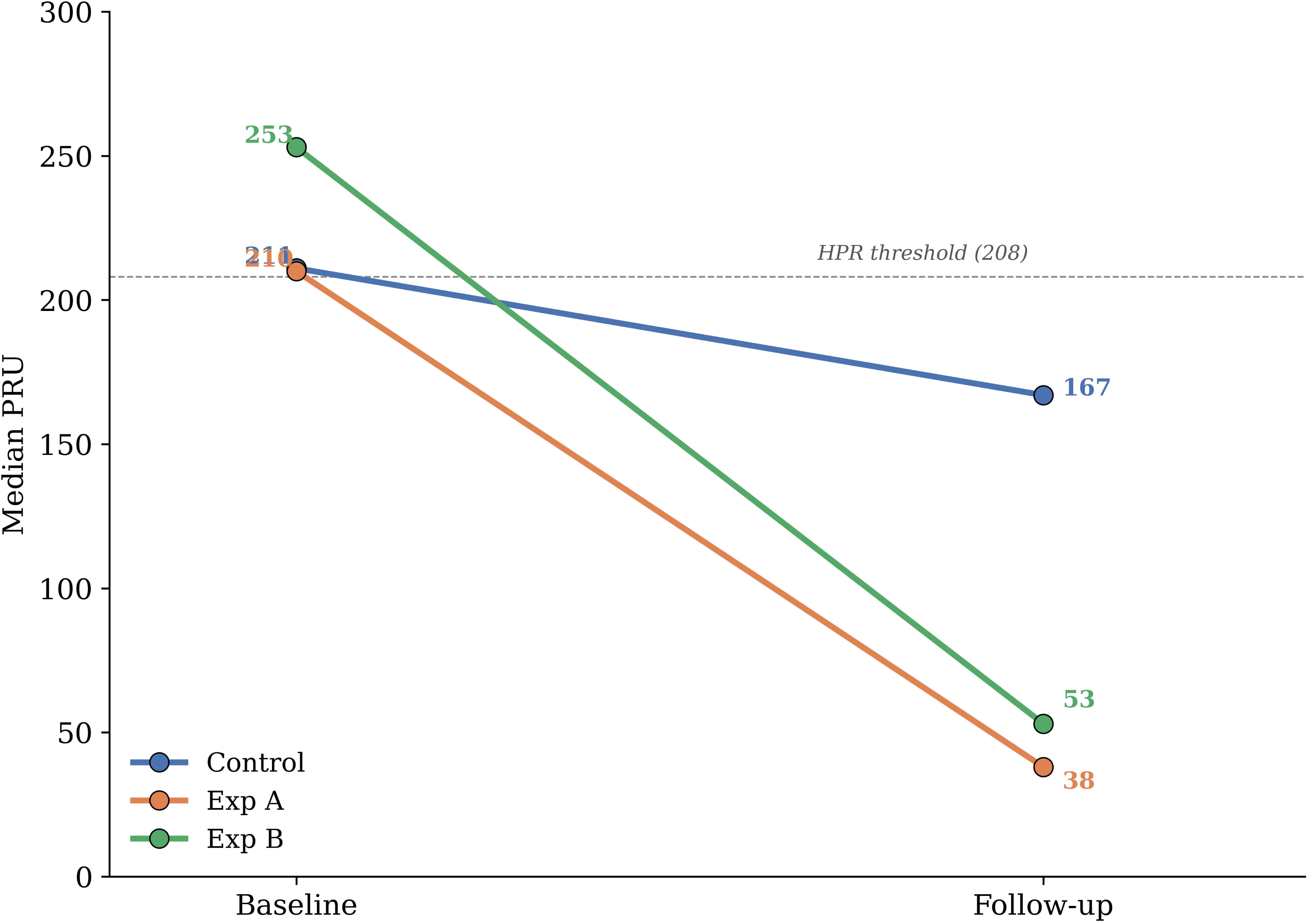
Individual and group P2Y_12_ reaction unit (PRU) values at baseline and follow-up. Each symbol represents one patient; horizontal bars indicate group medians; whiskers show interquartile range. PRU fell significantly within each arm from baseline (pre-loading dose) to follow-up (median 17 days post-PCI). Follow-up PRU was substantially lower in both ticagrelor arms than in Control. BID indicates twice daily; CCS, chronic coronary syndrome; DAPT, dual antiplatelet therapy; PCI, percutaneous coronary intervention; PRU, P2Y_12_ reaction units.

**Figure 3.**
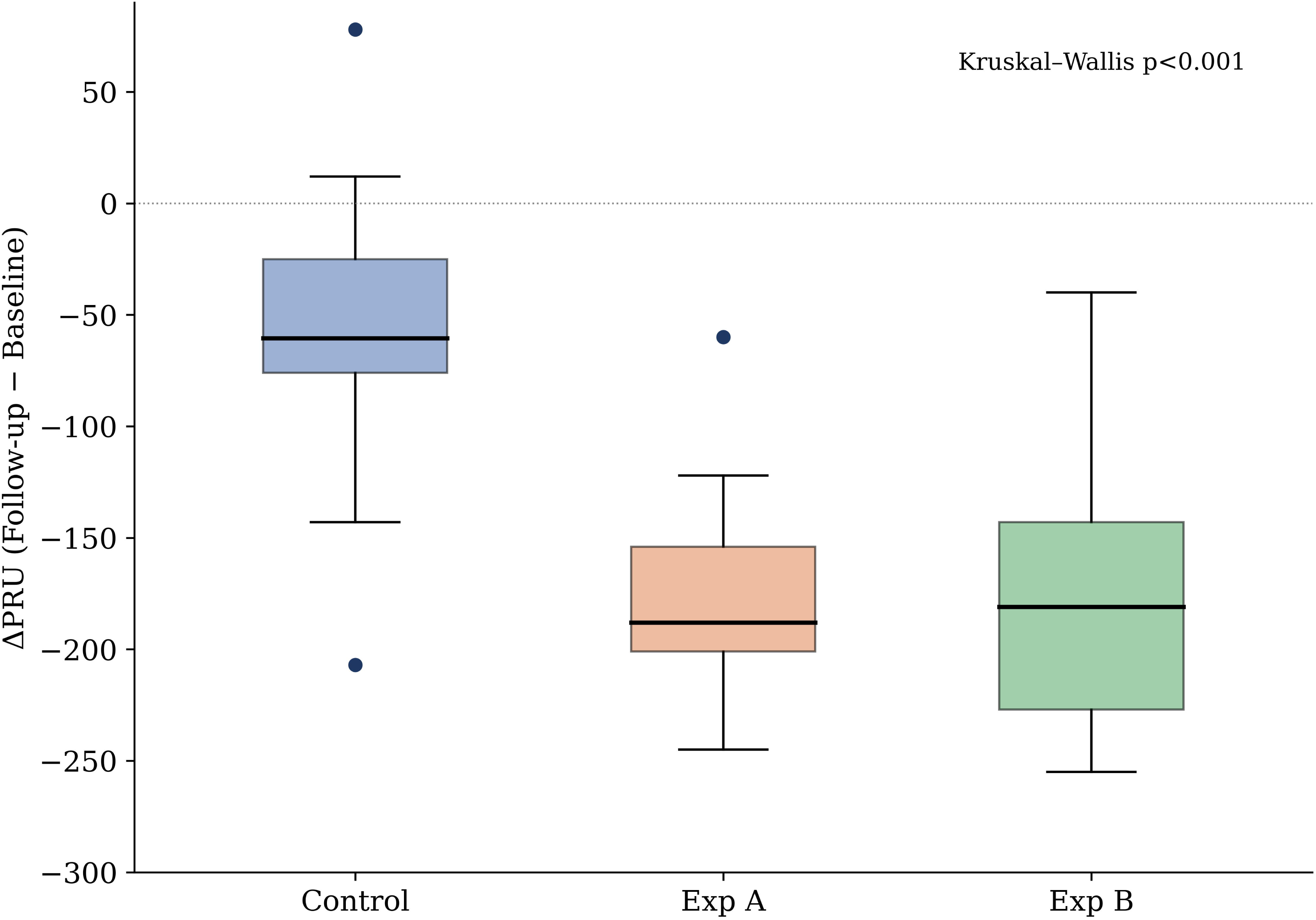
Between-group comparison of change in PRU (ΔPRU). Box plots show median and interquartile range; individual data points and whiskers are plotted separately. The overall between-group difference was significant (Kruskal–Wallis *P*<0.001); both ticagrelor arms showed markedly greater PRU reduction than Control, with no difference between Experimental A and B (*P*=0.772). Abbreviations as in Figure 2.

**Table 3.**
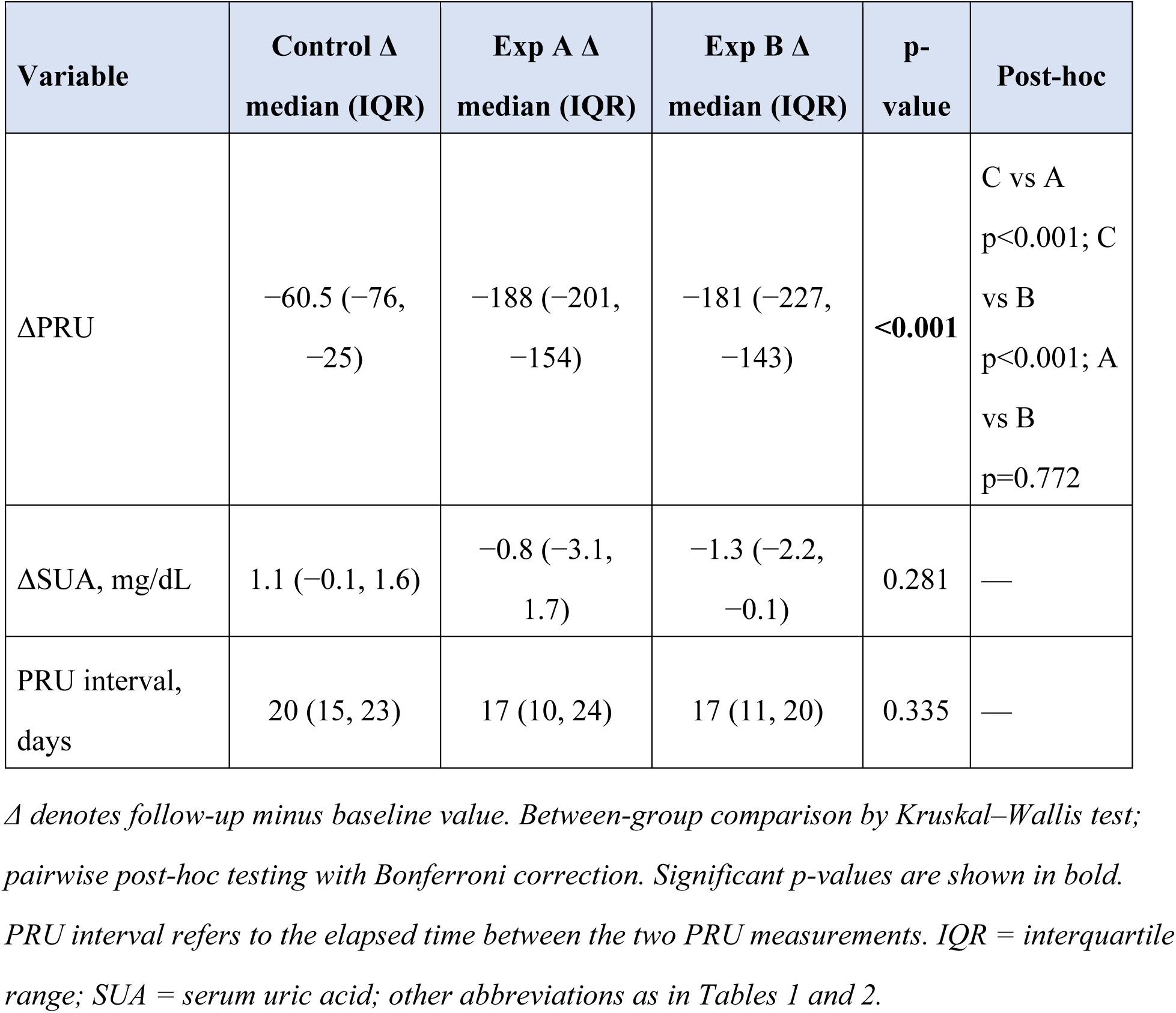
Between-group comparison of change scores (Δ) in clinical and laboratory parameters.

### Clinical outcomes

Over 12 months, MACE occurred in 0 of 18 Control patients, 0 of 19 Experimental A patients, and 1 of 17 Experimental B patients (5.9%; Figure 4). The single MACE was a target-vessel revascularization for in-stent restenosis of a right coronary artery chronic total occlusion in a patient with left main and triple-vessel disease and a markedly elevated lipoprotein(a) of 79.1 mg/dL, whose post-PCI PRU was 26. Any clinically relevant bleeding occurred in 0, 0, and 1 patient, respectively; the single bleeding event, in Experimental B, was a non-major gastrointestinal bleed. One Experimental A patient crossed over to aspirin plus clopidogrel for ticagrelor-related dyspnea; no other patient discontinued the assigned strategy. There were no episodes of stent thrombosis, no cardiac deaths, and no strokes (Table 4).

**Figure 4.**
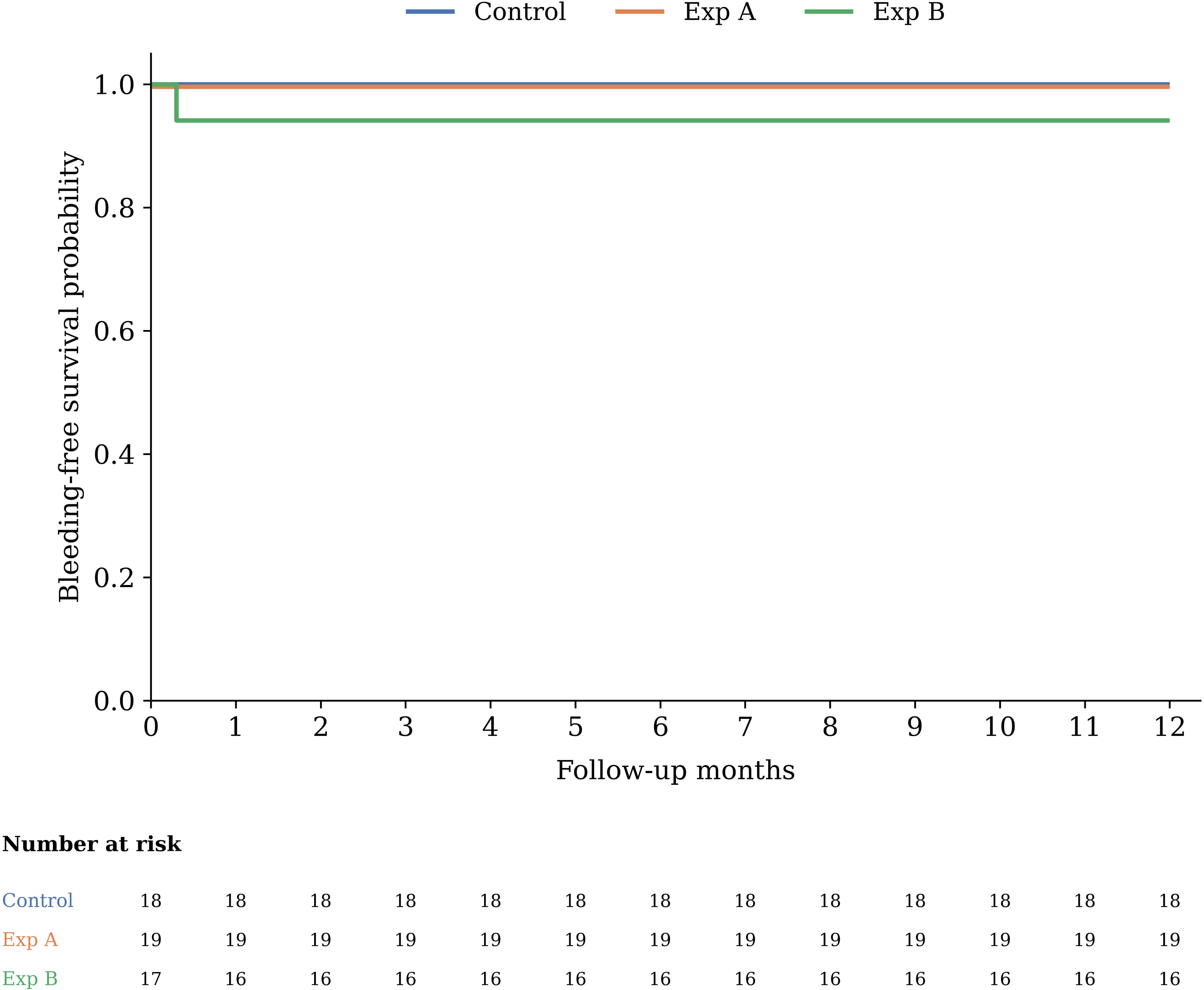
Clinically relevant bleeding-free survival. Kaplan–Meier estimates of freedom from any clinically relevant bleeding (Bleeding Academic Research Consortium criteria) over 12 months. A single non-major gastrointestinal bleeding event occurred in Experimental B; no bleeding events occurred in Control or Experimental A. The number at risk is shown below the plot. Abbreviations as in Figure 2.

**Figure 5.**
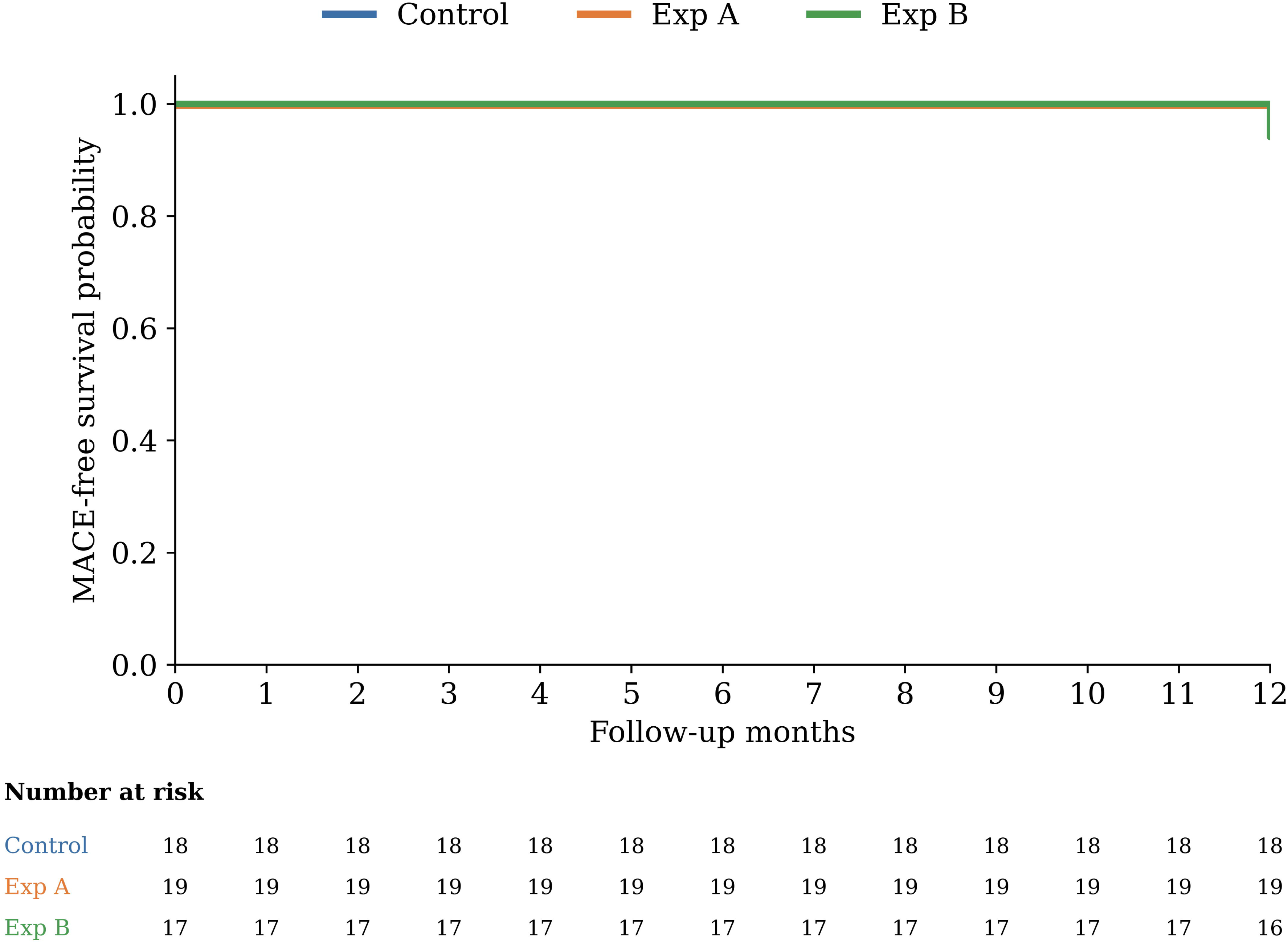
MACE-free survival. Kaplan–Meier estimates of freedom from major adverse cardiovascular events (MACE: a composite of all-cause death, target-vessel or target-lesion revascularization, and recurrent myocardial infarction) over 12 months. A single MACE occurred in Experimental B; no events occurred in Control or Experimental A. The number at risk is shown below the plot. MACE = major adverse cardiovascular events; other abbreviations as in Figure 2.

**Table 4.** Twelve-month composite clinical outcomes.

| Outcome at 12 months | Control<br>(n=18) | Exp A<br>(n=19) | Exp B<br>(n=17) | p-value |
| --- | --- | --- | --- | --- |
| MACE, n (%) | 0 (0.0) | 0 (0.0) | 1 (5.9) | 1.000† |
| Any clinically relevant bleeding, n (%) | 0 (0.0) | 0 (0.0) | 1 (5.9) | 1.000† |
| Crossover, n (%) | 0 (0.0) | 1 (5.3) | 0 (0.0) | — |

† Fisher's exact test. MACE was a composite of all-cause death, target-vessel or target-lesion revascularization, and recurrent myocardial infarction. Clinically relevant bleeding was classified per the Bleeding Academic Research Consortium criteria. Crossover refers to discontinuation of the assigned strategy during follow-up; the single crossover occurred in one Experimental A patient who was switched to aspirin plus clopidogrel for ticagrelor-related dyspnea.

## Discussion

Three findings emerge from this trial. First, half-dose ticagrelor produced an approximately three-fold greater reduction in PRU than aspirin plus clopidogrel — a disparity that is not subtle. Second, the pharmacodynamic effect of half-dose ticagrelor was identical whether given as DAPT or as monotherapy from the day of PCI: aspirin added no detectable platelet-inhibition benefit on top of ticagrelor 45 mg twice daily. Third, an aspirin-free strategy initiated at the index procedure was clinically tolerable in our cohort, with no stent thrombosis, no cardiac death, and only a single ticagrelor-related dyspnea crossover across the entire study.

These observations extend the prevailing direction of post-PCI antiplatelet research. TWILIGHT, TICO, T-PASS, and ULTIMATE-DAPT have collectively challenged the assumption that aspirin must be continued through the early post-stent period.^4–7,16^ Each of those trials, however, preserved a defined window of conventional DAPT before withdrawing aspirin and used ticagrelor at the standard 90 mg twice-daily dose. The present trial removes both constraints simultaneously: aspirin is omitted from hospital discharge and the maintenance dose is halved.

That the pharmacodynamic profile remains well within the therapeutic range under these conditions argues that the structural conservatism of contemporary post-PCI antithrombotic algorithms — multi-drug, multi-step, calibrated for the highest-risk population — may be unnecessary in the East-Asian CCS phenotype.^17–19^

The follow-up PRU values in our half-dose ticagrelor arms (medians 38 and 53) sit well below the conventional HPR cutoff of 208, and indeed within the range that some authors have associated with bleeding excess.^9–11,20^ Whether this signals overtreatment in East-Asian patients — and whether an even lower dose, or intermittent dosing, might achieve adequate ischemic protection with a tighter bleeding margin — is the natural next question.^12–14^

Several clinical observations merit comment despite the small sample size. No stent thrombosis occurred in either ticagrelor arm, including the 17 patients who left the catheterization laboratory on ticagrelor monotherapy. The single ticagrelor-related dyspnea crossover (one patient in Experimental A, 5.3%) is consistent with prior reports and reinforces standard counseling rather than constraining the strategy. The single MACE in the monotherapy arm is also instructive: it was a target-vessel revascularization for in-stent restenosis of a right coronary artery chronic total occlusion — not a thrombotic event — and it occurred despite near-maximal platelet inhibition (PRU 26) in a patient carrying a high atherosclerotic burden (left main and triple-vessel disease) and a markedly elevated lipoprotein(a) of 79.1 mg/dL. Lipoprotein(a) is an established driver of accelerated atherosclerosis and restenosis,^21^ and this case illustrates that residual ischemic risk in such patients is mediated by mechanisms that potent P2Y_12_ inhibition cannot address — consistent with the principle that half-dose ticagrelor effectively controls platelet-mediated risk while anatomical complexity and lipoprotein(a)-related risk require separate management strategies.

The procedural profile of the three arms also merits comment. Although randomization produced groups that were well matched for demographics, comorbidities, and laboratory variables, Experimental A had the most demanding anatomy and the heaviest interventional burden: it ranked numerically highest on every complexity measure recorded — ACC/AHA type B2/C lesions, multivessel and triple-vessel disease, and chronic total occlusion — and received significantly more stents (median 3 [IQR 2–4] vs. 2 [1–2] and 1 [1–4]; *P*=0.033) and a greater total stent length (91 [49–129] mm vs. 47.5 [30–73] mm and 33 [18–92] mm; *P*=0.023) than the Control and monotherapy arms. With only 18–19 patients per group, such imbalance is an expected consequence of small-sample randomization rather than evidence of selection bias. The greater stent count and total stented length in Experimental A also reflect a deliberate procedural philosophy: patients with multivessel disease who declined coronary artery bypass grafting after detailed shared decision-making underwent complete percutaneous revascularization, which necessarily increased the stent count and stented length in the arms that happened to contain more such patients. This imbalance has two implications. First, any comparison of clinical events across arms must be interpreted with caution, because the arm carrying the greatest ischemic and procedural risk was not the arm with the simplest regimen; the low and statistically indistinguishable event rates should therefore be regarded as hypothesis-generating only. Second, and reassuringly, the pharmacodynamic finding is strengthened rather than weakened by this imbalance: despite a substantially heavier stent burden, Experimental A achieved a PRU reduction essentially identical to that of the less complex monotherapy arm, indicating that the potent and uniform platelet inhibition of half-dose ticagrelor was independent of lesion complexity and stented length. A definitive trial should nonetheless stratify or adjust for lesion complexity and stent burden to ensure that any difference in ischemic or bleeding outcomes can be attributed to the antiplatelet strategy itself.

If reproduced in a larger trial, half-dose ticagrelor monotherapy initiated at PCI offers several concrete practical and pharmacological advantages: a single drug from day one; elimination of the well-documented gastrointestinal and hemorrhagic toxicity of chronic aspirin; dosing calibrated to East-Asian pharmacokinetics;^12^ a reduced periprocedural-bleeding burden for the not-infrequent patient who subsequently requires non-cardiac surgery;^22^ and removal of the predictable adherence inflection that occurs at the conventional six- or 12-month DAPT transition. A multicenter randomized trial, powered for non-inferiority against standard DAPT for ischemic endpoints and for superiority for clinically relevant bleeding, is the logical next step. Sub-questions of value would include whether patients with prior myocardial infarction — in whom long-term ticagrelor has shown benefit^23^ — and stable diabetic patients^24^ derive disproportionate benefit, and whether platelet-function-guided de-escalation^25,26^ adds value over a fixed half-dose protocol.

### Limitations

The trial is small, single-center, and open-label; clinical event rates are correspondingly low and all endpoint analyses must be regarded as exploratory. Twelve-month follow-up cannot yet address the long-term safety of the indefinite-continuation monotherapy arm. The findings are specific to an East-Asian CCS population and should not be extrapolated to ACS or to non-Asian cohorts without further data. PRU was used as a descriptive readout rather than as a treatment-guidance tool. Despite randomization, Experimental A received significantly more stents and a greater total stent length than the other arms — an imbalance that may confound the clinical-event comparison and underscores the need for caution in interpreting the exploratory outcome data. The pharmacodynamic signal, however, is large, internally consistent, and robust to the constraints imposed by sample size.

## Conclusions

In East-Asian patients with CCS undergoing PCI, half-dose ticagrelor produced markedly greater platelet inhibition than aspirin plus clopidogrel, and reached the same pharmacodynamic level whether given with or without aspirin from the day of the procedure. Aspirin-free half-dose ticagrelor monotherapy initiated at PCI is feasible and warrants definitive evaluation in an adequately powered randomized trial.

## Supporting information

https://drive.google.com/drive/u/0/folders/1KBMXOvbPQ9a8_y-p_b3Eff_sCchOhNr5

## Nonstandard Abbreviations and Acronyms

ACS: acute coronary syndrome
BARC: Bleeding Academic Research Consortium
CCS: chronic coronary syndrome
DAPT: dual antiplatelet therapy
HPR: high platelet reactivity
MACE: major adverse cardiovascular events
PCI: percutaneous coronary intervention
PRU: P2Y₁₂ reaction units.

## Acknowledgments

The authors thank the clinical research coordinators, catheterization laboratory staff, and outpatient nursing team at KSVGH for their dedicated support of this trial, and the patients who consented to participate. The authors also thank personnel at the Health Examination Center and the Department of Medical Education and Research of Kaohsiung Veterans General Hospital for providing information in response to inquiries and for assistance with data processing. Special thanks are extended to Cai-Sin Yao for substantial contributions to data processing and statistical analysis.

## Author Contributions

F.Y.K., M.C.W., and G.Y.M. conceived and designed the study. F.Y.K., E.S.L., T.H.Y., and H.T.T. enrolled patients and performed PCI procedures. F.Y.K. and M.C.W. supervised PRU measurements and data collection. C.S.Y. and R.I.C. performed the statistical analysis. F.Y.K. drafted the manuscript. All authors critically revised the manuscript for important intellectual content and approved the final version for submission.

## Data Availability Statement

The data underlying this article will be shared on reasonable request to the corresponding author.

