## Supplementary material for "Half-Dose Ticagrelor Monotherapy Versus Standard Dual Antiplatelet Therapy in Chronic Coronary Syndrome After Percutaneous Coronary Intervention: A Randomized Pilot Trial With PRU-Guided Pharmacodynamic Assessment": https://drive.google.com/drive/u/0/folders/1KBMXOvbPQ9a8_y-p_b3Eff_sCchOhNr5

**Contents:** Supplemental Methods (eligibility criteria; platelet-function assay protocol; endpoint definitions, including BARC bleeding criteria and MACE components); CONSORT 2010 checklist for randomized pilot and feasibility trials.

### Supplemental Methods

#### S1. Eligibility criteria

##### Inclusion criteria

- Age ≥ *[20]* years (*[confirm lower age limit applied]*).
- Chronic coronary syndrome (CCS), defined per the 2019 European Society of Cardiology guidelines.
- Scheduled for elective percutaneous coronary intervention (PCI) on the day after admission.
- Willing and able to provide written informed consent before randomization.

##### Exclusion criteria

- Acute coronary syndrome (ACS) at presentation.
- Prior intolerance to any study drug (aspirin, clopidogrel, or ticagrelor).
- Active bleeding, or Bleeding Academic Research Consortium (BARC) ≥ 3 bleeding within the preceding 3 months.
- New York Heart Association (NYHA) class IV heart failure.
- Platelet count < $100 \times 10^9$/L.
- Estimated glomerular filtration rate (eGFR) < 30 mL/min/1.73 $m^2$.
- Severe hepatic dysfunction (*[define criterion applied, e.g., Child–Pugh class C]*).
- Planned non-cardiac surgery within 12 months.
- Inability to provide informed consent.

**Note.** Patients in whom PCI was not performed at the index procedure were withdrawn from the per-protocol analysis.

#### S2. Platelet-function (PRU) assay protocol

**Device.** Platelet reactivity was measured with the VerifyNow P2Y12 point-of-care assay (*[manufacturer, e.g., Werfen/Accriva Diagnostics]*) and reported in $P2Y_{12}$ reaction units (PRU).

**Sampling.** Whole-blood samples were collected into *[specify tube, e.g., 3.2% sodium citrate]* and processed within *[specify maximum interval, e.g., 0.5–4 h]* of draw, per the manufacturer's instructions.

**Timing.** Samples were obtained at two time points: (i) at baseline, before the index loading dose; and (ii) at follow-up, a median of 17 days after PCI (*[state the protocol window, e.g., 14 ± 7 days]*).

**Thresholds.** High platelet reactivity (HPR) was pre-specified as PRU ≥ 208 and very low platelet reactivity as PRU < 85.

**Blinding/quality control.** Assays were performed by laboratory personnel blinded to treatment allocation. *[Add device calibration / quality-control procedures and the personnel/credentials, if to be reported.]*

#### S3. Endpoint definitions

**Primary (pharmacodynamic) endpoint.** Change in PRU (ΔPRU), calculated as follow-up PRU minus baseline PRU.

**Secondary (clinical) endpoints, assessed over 12 months.**

- **Major adverse cardiovascular events (MACE):** a composite of all-cause death, target-vessel or target-lesion revascularization, and recurrent myocardial infarction (MI). Component definitions:
- All-cause death — death from any cause.
- Myocardial infarction — *[per the Fourth Universal Definition of MI; confirm and cite]*.
- Target-vessel / target-lesion revascularization — clinically driven repeat revascularization of the target vessel/lesion (*[per Academic Research Consortium definitions; confirm]*).
- **Clinically relevant bleeding:** classified by the BARC criteria. *[Specify which BARC types were counted as "clinically relevant," e.g., BARC type ≥ 2.]*
- **Study-drug intolerance or crossover,** including ticagrelor-related dyspnea.

**BARC bleeding classification (reference).**

| BARC type | Definition |
|---|---|
| Type 0 | No bleeding. |
| Type 1 | Bleeding that is not actionable and does not cause the patient to seek evaluation. |
| Type 2 | Any overt, actionable sign of hemorrhage that does not meet type 3–5 but requires intervention, hospitalization, or prompts evaluation. |
| Type 3a | Overt bleeding plus hemoglobin drop of 3 to < 5 g/dL; or any transfusion with overt bleeding. |
| Type 3b | Overt bleeding plus hemoglobin drop ≥ 5 g/dL; cardiac tamponade; bleeding requiring surgical intervention or intravenous vasoactive agents. |
| Type 3c | Intracranial or intraocular bleeding compromising vision. |
| Type 4 | Coronary artery bypass grafting–related bleeding. |
| Type 5a | Probable fatal bleeding. |
| Type 5b | Definite fatal bleeding. |

*BARC indicates Bleeding Academic Research Consortium. Source: Mehran R, et al. Circulation. 2011;123:2736–2747 (cited in the main reference list).*

### CONSORT 2010 checklist for randomized pilot and feasibility trials

*Adapted from Eldridge SM, Chan CL, Campbell MJ, Bond CM, Hopewell S, Thabane L, Lancaster GA; PAFS consensus group. CONSORT 2010 statement: extension to randomised pilot and feasibility trials. BMJ. 2016;355:i5239. Page numbers refer to the current manuscript draft and should be confirmed against the final paginated version. Items flagged in brackets indicate where reporting should be added or clarified before submission.*

| Section / Topic | Item | Checklist item | Reported in (section, page) |
|---|---|---|---|
| **Title and abstract** | | | |
| | 1a | Identification as a pilot or feasibility randomised trial in the title | Title page (p.1) — "A Randomized Pilot Trial" |
| | 1b | Structured summary of pilot trial design, methods, results, and conclusions (for specific guidance see CONSORT abstract extension for pilot trials) | Abstract (p.2–3) |
| **Introduction** | | | |
| Background and objectives | 2a | Scientific background and explanation of rationale for future definitive trial, and reasons for randomised pilot trial | Introduction (p.5) |
| | 2b | Specific objectives or research questions for pilot trial | Introduction, final paragraph (p.5) |
| **Methods** | | | |
| Trial design | 3a | Description of pilot trial design (such as parallel, factorial) including allocation ratio | Methods – Study design and ethics (p.6) |
| | 3b | Important changes to methods after pilot trial commencement (such as eligibility criteria), with reasons | Not applicable — no important changes after commencement |
| Participants | 4a | Eligibility criteria for participants | Methods – Patients (p.6); Supplemental Methods S1 |
| | 4b | Settings and locations where the data were collected | Methods – Study design and ethics (p.6) |
| | 4c | How participants were identified and consented | Methods – Study design and ethics (p.6) — written informed consent before randomization |
| Interventions | 5 | The interventions for each group with sufficient details to allow replication, including how and when they were actually administered | Methods – Randomization and interventions (p.6–7) |
| Outcomes | 6a | Completely defined prespecified assessments or measurements to address each pilot trial objective specified in 2b, including how and when they were | Methods – Endpoints; Pharmacodynamic assessment (p.7–8); Supplemental |

| Section / Topic | Item | Checklist item | Reported in (section, page) |
|---|---|---|---|
| | | assessed | Methods S2–S3 |
| | 6b | Any changes to pilot trial assessments or measurements after the pilot trial commenced, with reasons | Not applicable — no changes |
| | 6c | If applicable, prespecified criteria used to judge whether, or how, to proceed with future definitive trial | Not prespecified — objective was effect-size estimation (Methods – Statistical analysis, p.8). [Consider adding explicit progression criteria.] |
| Sample size | 7a | Rationale for numbers in the pilot trial | Methods – Statistical analysis (p.8) |
| | 7b | When applicable, explanation of any interim analyses and stopping guidelines | Not applicable — no interim analyses or stopping guidelines |
| Randomisation – sequence generation | 8a | Method used to generate the random allocation sequence | Methods – Randomization and interventions (p.6) — computer-generated sequence |
| | 8b | Type of randomisation(s); details of any restriction (such as blocking and block size) | Methods (p.6) — 1:1:1 allocation. [Restriction/block size not specified — add if used.] |
| Allocation concealment mechanism | 9 | Mechanism used to implement the random allocation sequence (such as sequentially numbered containers), describing any steps taken to conceal the sequence until interventions were assigned | Not reported — [add the concealment mechanism]. |
| Implementation | 10 | Who generated the random allocation sequence, who enrolled participants, and who assigned participants to interventions | Partially reported — enrolment in Author Contributions (p.14). [Specify who generated the sequence and assigned participants.] |
| Blinding | 11a | Who was blinded after assignment to interventions (for example, participants, care providers, those assessing outcomes) and how | Methods – Randomization and interventions (p.6) — open-label; PRU assays by personnel blinded to allocation |
| | 11b | If relevant, description of the similarity of interventions | Not applicable — open-label trial |
| Statistical methods | 12 | Methods used to address each pilot trial objective whether qualitative or quantitative | Methods – Statistical analysis (p.8) |
| **Results** | | | |

| Section / Topic | Item | Checklist item | Reported in (section, page) |
|---|---|---|---|
| Participant flow (a diagram is strongly recommended) | 13a | For each group, the numbers of participants who were approached and/or assessed for eligibility, randomly assigned, received intended treatment, and were assessed for each objective | Results – Patient characteristics (p.8); Figure 1 (CONSORT flow) |
| | 13b | For each group, losses and exclusions after randomisation, together with reasons | Results – Patient characteristics (p.8); Figure 1 (7 did not undergo PCI; 1 crossover) |
| Recruitment | 14a | Dates defining the periods of recruitment and follow-up | Methods – Study design and ethics (p.6) — 1 Jan–31 Dec 2024; 12-month follow-up |
| | 14b | Why the pilot trial ended or was stopped | Trial ran to its planned enrolment/feasibility target; not stopped early (Methods, p.6/8). [State explicitly.] |
| Baseline data | 15 | A table showing baseline demographic and clinical characteristics for each group | Results – Patient characteristics (p.8); Table 1 |
| Numbers analysed | 16 | For each objective, number of participants (denominator) included in each analysis. If relevant, these numbers should be by randomised group | Results – Patient characteristics (p.8); Figure 1 (analysis population 18/19/17) |
| Outcomes and estimation | 17 | For each objective, results including expressions of uncertainty (such as 95% confidence interval) for any estimates. If relevant, these results should be by randomised group | Results – Pharmacodynamic outcomes (p.10); Table 3; Figures 2–3. [Medians with IQR reported; consider adding 95% CIs for effect-size estimates.] |
| Ancillary analyses | 18 | Results of any other analyses performed that could be used to inform the future definitive trial | Results – procedural burden; blood pressure and serum uric acid (p.10) |
| Harms | 19 | All important harms or unintended effects in each group (for specific guidance see CONSORT for harms) | Results – Clinical outcomes (p.10–11); Table 4; Figure 4 (MACE, bleeding) |
| | 19a | If relevant, other important unintended consequences | Results – Clinical outcomes (p.11) — single ticagrelor-related dyspnea crossover |
| **Discussion** | | | |
| Limitations | 20 | Pilot trial limitations, addressing sources of potential bias and remaining uncertainty about feasibility | Discussion – Limitations (p.13) |

| Section / Topic | Item | Checklist item | Reported in (section, page) |
|---|---|---|---|
| Generalisability | 21 | Generalisability (applicability) of pilot trial methods and findings to future definitive trial and other studies | Discussion – Limitations (p.13) — East-Asian CCS specificity |
| Interpretation | 22 | Interpretation consistent with pilot trial objectives and findings, balancing potential benefits and harms, and considering other relevant evidence | Discussion (p.11–13) |
| | 22a | Implications for progression from pilot to future definitive trial, including any proposed amendments | Discussion; Conclusions (p.13–14) |
| **Other information** | | | |
| Registration | 23 | Registration number for pilot trial and name of trial registry | Methods – Study design and ethics (p.6); Abstract – Clinical Trial Registration (p.3) — ClinicalTrials.gov NCT07622056 |
| Protocol | 24 | Where the pilot trial protocol can be accessed, if available | Not reported — [state where the protocol can be accessed, or that it is available from the corresponding author]. |
| Funding | 25 | Sources of funding and other support (such as supply of drugs), role of funders | Sources of Funding (p.15) |
| | 26 | Ethical approval or approval by research review committee, confirmed with reference number | Methods – Study design and ethics (p.6) — IRB approval KSVGH 23-CT12-07 |
